# Glaucoma and Diabetes Mellitus: A Comparative Evaluation of Comorbid Impact on Tear Quantity among Patients in Owerri, Imo State, Nigeria

**DOI:** 10.64898/2026.08.30.26361782

**Authors:** Chigozie Mary Chukwuoha, Emmanuel C. Esenwah, Nwakaego C. Ikoro, Godwin-Ovenseri-Ogbomo, Young Christian Azuamah, Nkiru Euphresia Odimegwu, Anthony U. Megwas, Genevieve Ugwoke, Jacqueline E. Obioma-Elemba, Yadirichukwu Eronini, Ezinne Chinenye Nkeremuzor

## Abstract

**Objective:** Glaucoma is a chronic disorder that impairs ocular health and may exacerbate ocular surface disease leading to tear film instability, dry eye symptoms and decreased quality of life. This study compared changes in tear quantity among glaucoma subjects living with and without diabetes mellitus, attending an eye clinic in Nigeria.

**Methods:** A comparative cross sectional research design was used. 157 subjects which comprised 74 glaucoma subjects living with diabetes mellitus and 83 glaucoma subjects living without diabetes mellitus participated in the study. Tear quantity assessment included the Schirmer I test and tear meniscus height (TMH) measurement. Descriptive statistics, independent samples t-test and Chi-square test were used to examine the data at 0.05 level of significance.

**Results:** Glaucoma subjects living with diabetes mellitus showed substantially decreased tear production (11.4 ± 6.8 mm) compared with glaucoma subjects living without diabetes mellitus (19.6 ± 9.6 mm; p < 0.001). Tear meniscus height in glaucoma subjects living with diabetes mellitus (0.8 ± 0.3 mm) was significantly greater than in subjects living without diabetes mellitus (0.7 ± 0.3 mm; p = 0.034).

**Conclusion:** Diabetes mellitus dramatically deteriorates the ocular surface function in glaucoma subjects by decreasing tear production, altering the tear meniscus height and increasing the severity of ocular surface symptoms. Routine glaucoma care, especially in patients with diabetes mellitus, should include a full ocular surface evaluation including Schirmer I test, TBUT, TMH, and OSDI assessment to allow early detection and management of ocular surface disease, better treatment adherence, and improved visual outcomes.

## 1. Introduction

The cornea, conjunctiva, tear film, lacrimal gland, nasolacrimal system and the eyelids constitute the ocular surface known as a complex and unified system.^1–4^ The ocular surface also comprise the surface and epithelial gland of the cornea, conjunctiva, lacrimal gland, accessory lacrimal glands, meibomian glands and their apical (tears) and basal (connective tissue) origins, the eyelashes with their related glands of Moll and Zeis, as well as features of the eyelids associated with blink and the nasolacrimal duct. Together, these features are linked through a continuous epithelium, as well as the nervous, vascular, immune and endocrine systems.^2,4–6^

The primary function of the Ocular Surface System is to maintain and protect the smooth refractive surface of the cornea.^7^ This function depends on the coordinated activities of structurally continuous epithelial tissues derived from the surface ectoderm. The corneal and conjunctival epithelia are anatomically continuous with the ductal epithelium of the lacrimal glands, accessory lacrimal glands, meibomian glands and the nasolacrimal drainage system. Such continuity permits communication among these tissues through cytokines, gap junctions and other cellular signaling mechanisms that regulate ocular surface maintenance and repair. In addition, each component contributes specific constituents to the tear film. The corneal and conjunctival epithelia synthesize hydrophilic mucins that facilitate tear adhesion, whereas the lacrimal and accessory lacrimal glands secrete the aqueous component together with numerous protective proteins. The meibomian glands produce the superficial lipid layer, which minimizes tear evaporation, while the nasolacrimal drainage system regulates tear outflow to maintain an appropriate balance between tear secretion and drainage.^4,5^

Tears have a vital function to protect and lubricate the ocular surface. Tear production, distribution and clearance is tightly regulated by the lacrimal functional unit (LFU) to meet ocular surface demands. The tear film consists of an aqueous-mucin layer, containing fluid and soluble factors produced by the lacrimal glands and mucin secreted by the goblet cells that are covered by a lipid layer. Tear production (about 1-2 microliters per minute, total volume 6 microliters, 16% turnover per minute), evaporation, absorption and drainage are responsible for dynamic balance of the preocular tear film.^8–10^

The World Health Organization (2018) globally identifies glaucoma as the major cause of irreversible blindness. It accounts for 8% of all blindness, affecting an estimated 3.12 million blind people.^11^ A review on the relevant population-based surveys of glaucoma, and of blindness and visual impairment in sub-Saharan Africa indicate that glaucoma affects about 4 % of adults aged 40 years and above and accounts for 15 % of blindness.^11–13^ Africa is the region with the highest incidence and prevalence of glaucoma, most of which is open-angle glaucoma (OAG).^11,14^ Although the pathogenesis of glaucoma is not fully understood, the incidence of glaucoma increases with age, the patients often have numerous comorbidities and use various medications.^15^ Moreover, increased intraocular pressure (IOP) is the most significant risk factor in glaucoma.^16^

Long term topical glaucoma therapy has been associated with reduced density of goblet cells and squamous metaplasia of the conjunctival epithelium,^17,18^ dysfunction of meibomian glands, conjunctival and corneal desquamation (Di Staso et al., 2018) and overexpression of proinflammatory cytokines.^19^ Significant loss of goblet cells, which can cause dry eye, inflammation, and fibrosis, was observed in animal and human models.^17,18^ As a consequence of the inflammatory changes, chronic use of IOP-lowering medications can also affect bleb scarring in filtration surgery, since it is a risk factor for conjunctival fibrosis which can ultimately result in failure of trabeculectomy.^20^

Investigations on the coexistence of glaucoma and ocular surface disease (OSD) estimated prevalence of OSD to vary between 5% and 30% in the general population, while noting that it may increase up to 50% in glaucoma patients under medical treatment.^2^ Several risk factors such as aging, hormone imbalance, systemic comorbidities, systemic medications and environmental exposure are frequently associated with the chronic use of IOP-lowering eye drops, triggering proinflammatory responses and ocular surface dysfunction in patients with glaucoma.^21^

Diabetes mellitus (DM) is a group of metabolic diseases or metabolic syndrome where there is hyperglycemia as a result of defect in the insulin secretion and/or action. In diabetics, chronic hyperglycemia is often associated with long-term damage, dysfunction, and/or failure of various organs, especially the eyes, kidneys, nerves and blood vessels. DM is a serious and increasingly prevalent health problem worldwide due to sedating lifestyle & population ageing.^22^ Hyperglycemia, caused by insulin production, insulin action or both is the hallmark of diabetes mellitus, manifesting a group of diabetic complications.

Studies have reported that the risk of glaucoma increased in diabetic cases with strong evidence in support of a positive association between glaucoma and diabetes.^23^ Ocular surface disease is also reported as one of the major side effects of topical anti-glaucoma medications. This study therefore aimed to compare the changes in the tear quantity among glaucoma subjects living with and without diabetes by assessing the tear production and tear meniscus height among subjects included in the study.

Specific research questions addressed by this study included: (i) What is the level of tear production among glaucoma subjects living with and without diabetes mellitus in Owerri, Imo State, Nigeria? (ii) What is the TMH among glaucoma subjects living with and without diabetes mellitus in Owerri, Imo State, Nigeria? (i) What is the difference in the level of tear production among glaucoma subjects living with and without diabetes mellitus in Owerri, Imo State, Nigeria? (ii) What is the difference in the TMH among glaucoma subjects living with and without diabetes mellitus in Owerri, Imo State, Nigeria? It was hypothesized as well that there was no difference in the tear production and tear meniscus height among glaucoma subjects living with and without diabetes mellitus in Owerri, Imo State, Nigeria.

## 2. Material and Methods

### 2.1 Research design

Data used in study were drawn from a larger clinic-based cross-sectional study investing the comparative assessment of ocular surface changes among glaucoma subjects living with and without diabetes mellitus in owerri, Imo State, Nigeria.

### 2.2 Population of study

The study population comprised 157 subjects who gave informed consent and met with the inclusion criteria for this study, diagnosed with glaucoma and living with or without diabetes, who visited Mercy Eye Clinic, Owerri, during the study period.

### 2.3 Inclusion criteria

Subjects diagnosed of open-angle glaucoma, who were one or more topical antiglaucoma medication, including those living with diabetes mellitus as a comorbidity were included in the study.

### 2.4 Exclusion criteria

Subjects with history of ocular trauma, blepharitis, entropion, any ocular surgery or laser procedure within the last 6 months, any corneal pathologies, contact lens wearers, chronic use of other topical medication. Others are systemic diseases such as rheumatoid arthritis, Sjogren’s syndrome, thyroid disease, lupus, dry climate smoke and continuous computer screen users.

### 2.5 Procedure for data collection

First, a comprehensive ocular history was obtained to access data such as age, gender, time since glaucoma and diabetes diagnosis, use of topical antiglaucoma medication, previous ocular surgeries and then, ocular examination.

### 2.6 Tear production

For assessment of tear quantity, Schirmer I test and the tear meniscus height (TMH) were used. Schirmer test I is invasive while Meniscometry is a noninvasive way to assess tear volume indirectly by measuring the tear meniscus radius.

#### 2.6.1 Schirmer I Test

The Schirmer I test was used to evaluate aqueous-deficient dry eye by measuring basal tear secretion in response to conjunctival stimulation. It was performed by first instlling topical anesthetic (Amethocaine) to reduce sensitivity. Then a strip of Whatman #41 paper was placed at the junction of the middle and lateral thirds of the lower eyelid of each participant’s eye, to minimize irritation to the cornea. The participants were asked to look forward and blink normally while the strip was held in place for 5 minutes. After 5 minutes, the strips was examined and the length of wetting was recorded in millimeters. The results were graded as follows:

|  |  |  |
| --- | --- | --- |
| <5 | - | Severe dry eye |
| 5-10 mm | - | Mild to moderate dry eye |
| >10 mm | - | Normal |

With the use of topical anesthetic, values less than 5mm were considered diagnostic of aqueous tear deficiency and values between 5 to 10mm were considered suggestive.

#### 2.6.2 Tear meniscus height

Each participant was administered 2 drops of 1% sodium fluorescein solution. This was precisely instilled into the lower conjunctival sac utilizing a bottle dropper, ensuring accurate and consistent dosing for all participants. Following this instillation, the TMH was measured exactly 3 min after the fluorescein solution administration under wide illumination using a cobalt blue filter on a slit lamp biomiscroscope. This timing was crucial to allow the solution to disperse and interact with the eye’s surface properly. TMH is reduced in aqueous-deficient dry eye as indicated by a reduced height and radius of curvature. The values were graded as follows:

|  |  |  |
| --- | --- | --- |
| <1mm | - | grade 1 |
| >1mm | - | grade 2 |

### 2.7 Statistical analysis

The IBM Statistical Package for the Social Sciences (SPSS) version 31 was used for data analysis. Quantitative variables were described using mean and standard deviation (SD) for continuous variables whereas categorical variables were presented using frequencies and percentages. Independent sample t-test was used to determine the difference between the two groups. The chi-square test was used to analyze categorical variables. Correlation analysis was used to assess the relationship between the variables in the study. A significance level (*p value*) of ≤ 0.05 was considered statistically significant for all variables in the study.

### 2.8 Ethical Approval/Informed Consent

Ethical Approval was obtained from the Ethics Committee, School of Health Technology, Federal University of Technology, Owerri, to carry out this study after review, on the 8^th^ of July, 2025. No specific protocol number was assigned by the committee. Informed consent was also gotten from subjects before inclusion in the study.

## 3. Results

A total of 157 subjects comprising 48 (30.6%) males and 109 (69.4%) females made up the study population. The mean age of all subjects included in the study was 62.9±11.1. Among the male subjects, the mean age was 64.1±11.0 yeas, while among the female subjects, the mean age was 62.3±11.1. The age groups ranged from 25 to 94 years, with a frequency of 42.0% (45.8% males, 40.4% females) among those in age group 65-74 years which made up the highest population of the study. This was followed by 55-64 years age group, making up 29.9% 9 (31.3% males, 29.4% females), 45-54 years making up 11.5% (6.3% males, 13.8% females), 75-84 years making up 8.3% (4.2% males, 10.1% females), 35-44 years making up 5.1% (8.3% males, 3.7% females) and 25-34 years making up 1.9% (0.0% males, 2.8% females). The least population was seen among the 85-94 years age group which up 1.3 % (4.2% males, 0.0% females) of the study population.

**Figure 1:**
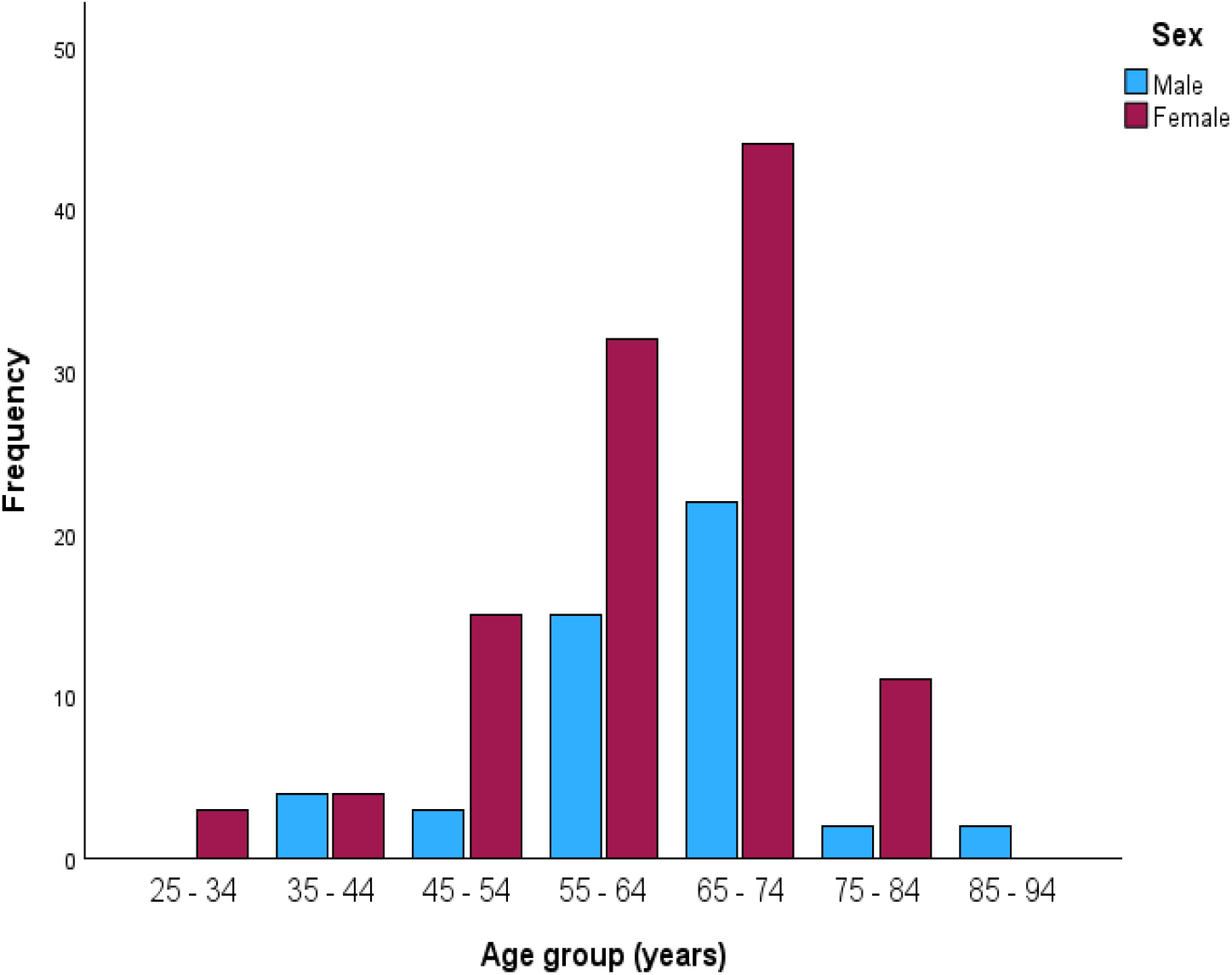
Overall Age and Sex distribution of glaucoma subjects living with and without DM in Owerri

### Tear Production

Table 1 below shows the level of tear production among glaucoma subjects living with and without DM in Owerri. The mean level of tear production among all subjects incuded in this study was 15.7± 9.3. For glaucoma subjects living with DM in Owerri, the mean tear production was 11.4± 6.8, while for glaucoma subjects living without DM, the mean tear production was 19.6±9.6.

**Table 1:** Level of Tear Production among Glaucoma subjects living with and without DM in Owerri.

| **Tear Production(mm)** | **Glaucoma subjects living with DM** | **%** | **Glaucoma subjects living without DM** | **%** | **Total** | **% Total** |
| --- | --- | --- | --- | --- | --- | --- |
| **>10 (Normal)** | 40 | 54.1 | 74 | 89.2 | 114 | 72.6 |
| **6-10 (mild-moderate)** | 19 | 25.7 | 3 | 3.6 | 22 | 14.0 |
| **1-5 ( Severe)** | 15 | 20.3 | 6 | 7.2 | 21 | 13.4 |
| **Total** | **74** | **100** | **83** | **100** | **157** | **100** |

Overall, 72.6% of the subjects in this study had level of tear production above 10mm. This was followed by 14.0% of the population with level of tear production between 6-10mm. The least level of tear production was seen among 13.4% of the study population, who had level of tear production n between 1-5mm.

Among the population of glaucoma subjects living with DM in Owerri, 54.1% had level of tear production above 10mm. This was followed by 25.7% of subjects in this group, with level of tear production between 6-10mm, while the least level of tear production was seen among 20.3% of glaucoma subjects living with DM, who had level of tear production between 1-5mm.

For glaucoma subjects living without diabetes mellitus in Owerri, 89.2% had level of tear production above 10mm. This was followed by 3.6% of subjects in this group, had level of tear production between 6-10mm, while the least occurence was seen among 7.2% of subjects in this group, who had level of tear production between 1-5mm.

### Tear Menicus Height

Table 2 below shows the TMH among glaucoma subjects living with and without DM in Owerri. The mean TMH among all subjects incuded in this study was 0.7± 0.3mm. For glaucoma subjects living with DM in Owerri, the mean TMH was 0.8± 0.3, while for glaucoma subjects living without DM, the mean TMH was 0.7±0.3.

**Table 2:**
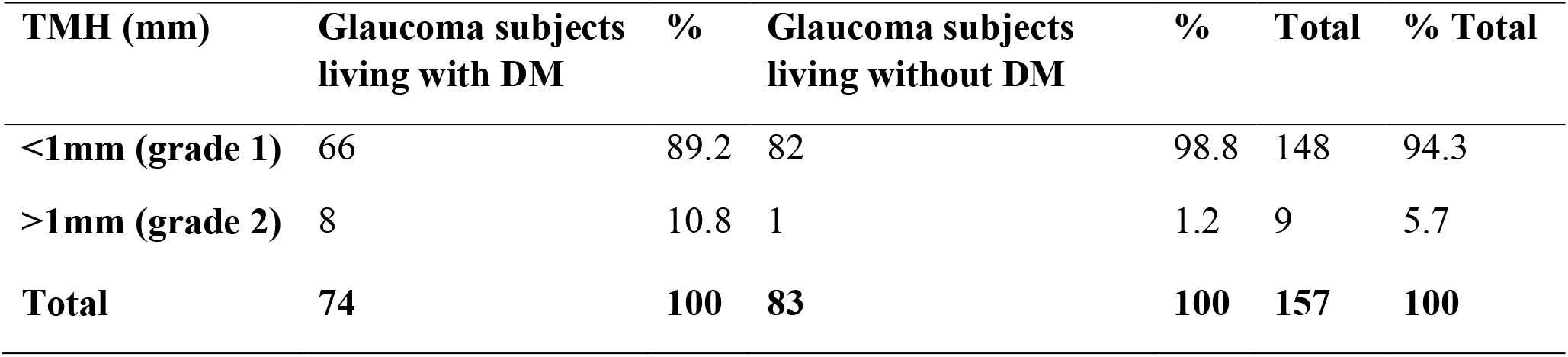
TMH among glaucoma subjects living with and without diabetes mellitus in Owerri.

Overall, 5.7% of the subjects in this study had TMH above 1mm while 94.3% had TMH below 1mm. Among the population of glaucoma subjects living with DM in Owerri, 10.8% had TMH above 1mm while 89.2% had TMH below 1mm. For glaucoma subjects living without DM in Owerri, 1.2% had TMH above 1mm while 98.8% of subjects in this group, had TMH below 1mm.

### Comparative effect of Glaucoma and Comorbid Diabetes Mellitus on Tear Production

Table 3 below shows the summary of data analysis for difference in the level of tear production among glaucoma subjects living with and without DM in Owerri, while testing the first hypothesis, indicating the *p-value* and Chi-square (χ^2^) results.

**Table 3:** Summary of data analysis for difference in the level of tear production among Glaucoma subjects living with and without Diabetes Mellitus in Owerri.

| Tear Production |  |  |
| --- | --- | --- |
| Glaucoma subjects living with DM | Glaucoma subjects living without DM | *P-value* |
| 74 | 83 | <.001 |

Chi square test $\chi^2 = 42.086$ $p < .001$

**Table 4:** Summary of data analysis for difference in the Tear Meniscus Height among Glaucoma subjects living with and without Diabetes Mellitus in Owerri.

| Tear Meniscus Height |  |  |
| --- | --- | --- |
| Glaucoma subjects living with DM | Glaucoma subjects living without DM | *P-value* |
| 74 | 83 | .034 |

Chi square $\chi^2 = 6.68$ , $p = .010$

At 0.05 significance level, the independent sample t-test showed a statistically significant difference (*p* < 0.001) in the mean tear production between glaucoma subjects living with and without DM in Owerri. Hence, the study observed a difference in the tear production among glaucoma subjects living with and without DM in Owerri. In terms of grading, the chi square analysis showed a statistically significant relationship between diabetes status and grading of tear production, χ^2^ = 42.086, *p* < .001. Hence, diabetes status was significantly associated with the measured level and grading of tear production among the glaucoma subjects included in the study.

### Comparative Impact of Glaucoma and Comorbid Diabetes Mellitus on Tear Meniscus Height

The independent sample *t*-test at 0.05 significance level showed a statistically significant difference (*p* = 0.034) in the mean TMH between glaucoma subjects living with and without DM in Owerri. Hence, the study observed a difference in the TMH among glaucoma subjects living with and without DM in Owerri. In terms of grading, the chi square analysis showed a statistically significant relationship between diabetes status and grading of TMH, χ^2^ = 6.68, *p* = .010. Hence, diabetes status was significantly associated with the measured level and grading of TMH among the glaucoma subjects included in the study.

## 4. Discussion

The majority of subjects in the 65 to 74 years age group is consistent with previous epidemiologic findings showing the prevalence of glaucoma increases markedly with increasing age. The prevalence of glaucoma increases progressively beyond the age of 50 years, and is greatest in those aged 70 years and above (Shan et al., 2024). Glaucoma is mostly burdening on the adult population aged 65 years and above due to the age-related degeneration of retinal ganglion cells, increased susceptibility of the optic nerve, impaired outflow of aqueous humour and cumulative exposure to systemic and ocular risk factors.^24,25^ Recent population estimates have reported a steep increase of the prevalence of glaucoma beyond the age of 60 years with the highest share of afflicted cases being in the age group ≥65 years, highlighting ageing as the most important demographic predictor of glaucoma.

The present findings also lend support to the landmark Barbados Eye Studies, among people of African descent, which showed a progressive increase in the incidence of glaucoma with age, with those aged 70 years and above having the highest risk of developing open-angle glaucoma.^26,27^ These studies share some relationship with the present study as the research population is comparable to the African ancestry of the individuals in the present study.

Tear production was assessed using Schirmer I test with anaesthesia. The study revealed glaucoma subjects living with DM had lower mean tear production (11.4 ± 6.8 mm) than glaucoma subjects living without DM in Owerri (19.6 ± 9.6 mm). Overall, 72.6% of the subjects exhibited normal tear production (>10 mm). This implies that aqueous tear production was not common among all subjects. This finding is particularly noteworthy and demonstrates the fact that adequate tear production does not rule out the presence of ocular surface disease symptoms as the tear film is maintained by the combined effort the mucus, lipid and aqueous layers. Hence, ocular surface disease or a compromise in the integrity of the ocular surface can still be observed among glaucoma subjects despite normal levels of tear production.

A previous study which assessed tear production with the schirmer test also reported normal level of tear production in majority of the subjects, ^18^ although a higher prevalence was seen in the present study. This may be attributable to a higher sample size used in the present study as compared to the previous study. Some other study demonstrated that there was no significant distinction in tear production with schirmer test result among participants despite increased OSDI scores and higher level of corneal staining.^28^ This also gives credence to the finding that clinically, ocular surface disease in glaucoma is not ruled out despite normal levels of tear production.

However, only 54.1% of glaucoma participants with DM had normal tear production compared 89.2% of non-diabetes subjects. Besides, decreased tear formation (≤10 mm) was reported in 45.9% of the glaucoma subjects living with DM against 10.8% of the non-DM glaucoma subjects. These results show that diabetes mellitus significantly impairs aqueous tear secretion in glaucoma subjects. The modest reduction in tear production did not differ significantly with some other study ^29^ which reported that changes in Schirmer test scores were not as evident as other ocular surface measures and continuous topical glaucoma treatment significantly impacts tear production and ocular surface integrity.

A comparative study among diabetic and non-diabetic volunteers in Nigeria reported diabetic subjects reported substantially lower Schirmer I test values as compared to non-diabetic controls, indicating compromised tear secretion.^30^ Another study in Western India,^31^ which assesed dry eye condition in type 2 diabetes mellitus patients using Schirmer I test, tear break-up time, fluorescein staining and OSDI also revealed that approximately 43.8% of diabetes individuals had dry eye condition with considerably worse tear function in diabetic retinopathy patients. The prevalent level of decreased tear production seen among glaucoma subjects with diabetes in the present study (45.9%) is similar to that described by the report these previous studies, indicating that diabetic mellitus no matter the geographic location, substantially reduces lacrimal gland function..^31^ The increased ocular surface effects observed in the current study may account for the greater burden of long-term topical antiglaucoma medications in chronic glaucoma.

Also, the findings of this study are consistent with those of another study in Ethiopia which noted a significant frequency in individuals with glaucoma and observed that extensive duration of therapy, use of multiple drugs and eye drops containing preservative, are important indicators for ocular surface disease.^32^ Although the prevalence reported was greater than the prevalence obtained in the present study, the variance is presumably due to different diagnostic criteria. However, both investigations indicate glaucoma subjects are more sensitive to ocular surface disease in long-term medicinal treatment.

While changes in Schirmer test scores were not as evident as other ocular surface measures in a previous study, it was reported that continuous topical glaucoma treatment significantly impacts tear production and ocular surface integrity.^31^ This is in line with the result of this study, that glaucoma subjects living without diabetes mellitus, showed modest reduction in tear production but those living with diabetes mellitus appeared to have increase in the alterations of the ocular surface. A similar evidence was provided by a systematic review and meta-analysis of 59 studies which revealed that persons with diabetes mellitus had substantially lower Schirmer I test scores than non-diabetic controls, suggesting a modest decrease in basal tear production associated with diabetes. The authors reported that the findings suggest an impairment of lacrimal gland function and tear film equilibrium in diabetes mellitus. It was also observed that inadequate glycaemic control worsened tear production, showing the role of persistent hyperglycemia in lacrimal gland dysfunction, corneal nerve damage and inflammation of the ocular surface,^33^ These mechanisms offer a physiologic basis for the dramatically decreased tear production reported in glaucoma subjects living with diabetes in the present investigation.

Hence, the present result suggest that diabetes mellitus has an additive detrimental effect on tear production in glaucoma sufferers. Chronic topical glaucoma treatments also contribute to ocular surface disease in both groups, the presence of diabetes considerably aggravates lacrimal gland function leading to a dramatically reduced tear production and a greater prevalence of aqueous-deficient dry eye. These findings stress the significance of frequent ocular surface screening in glaucoma patients, particularly in individuals with diabetes mellitus, to allow early intervention, treat compliance, ocular comfort and long-term visual result.

The overall mean Tear Meniscus Height (TMH) was 0.7±0.3 mm. Glaucoma subjects living with diabetes mellitus had a mean TMH somewhat greater (0.8 ± 0.3 mm) than those living without diabetes mellitus (0.7 ± 0.3 mm). In addition, the majority of the subjects (94.3%) had a TMH of less than 1 mm (grade 1) and just 5.7% had a TMH of more than 1 mm (grade 2). Glaucoma subjects living with diabetes mellitus had TMH < 1 mm in 89.2% of cases, while in glaucoma subjects living without diabetes mellitus it was 98.8%. The data reveal that the presence of decreased tear meniscus height was widespread in the study sample regardless of diabetes status, which may imply a diminishment of the tear reservoir in the glaucoma population.

The lower TMH among subjects in this study indicates reduced tear volume and possible ocular surface damage, which suggests that glaucoma, in addition to the chronic use of topical anti-glaucoma medications, may play a significant role in reducing the volume of the tears, which is further aggravated by DM as a comorbidity. This result is consistent with similar reports by previous studies^34,35,36^ with evidence that glaucoma subjects, especially those exposed to glaucoma treatment had lower TMH.

These findings are similar to those reported in a comprehensive review and meta-analysis of 24 observational studies on tear film changes in individuals with type 2 diabetes mellitus. The research found that diabetes patients usually had a decreased TMH compared to healthy controls, although the degree of reduction differed between studies. These variations were thought to be due to chronic diabetes-related changes in tear secretion and ocular surface homeostasis.^37^ Although the mean TMH in the present study was slightly higher for glaucoma subjects who lived with diabetes than for subjects who did not, the dominance of TMH values less than 1 mm in both groups is in line with the overall evidence that both glaucoma and diabetes are associated with compromised tear film dynamics.

The present finding can also be compared with the findings of Baek et al.^38^ who evaluated tear meniscus height using Fourier-domain anterior segment optical coherence tomography in individuals with type 2 diabetes mellitus. The study found that diabetic patients had considerably lower Tear Meniscus Height compared to healthy controls, with TMH decreasing gradually with the severity of diabetic retinopathy. The authors found that diabetes mellitus significantly impacted on tear volume and is involved in ocular surface disease. While the current study was conducted on glaucoma patients, not healthy controls, the majority of lower TMH values in the sample also indicates reduced tear volume in those with chronic ocular illness.

Nonetheless, the findings of the present study show that there is lower TMH among subjects with glaucoma in Owerri irrespective of diabetes status, which indicates reduced tear volume and possible ocular surface damage. There was a slightly higher mean TMH value in diabetic glaucoma subjects than in non-diabetic glaucoma subjects, but the great majority of values were below 1 mm in both groups, suggesting that glaucoma per se, in addition to the chronic use of topical anti-glaucoma medications, may play a significant role in reducing the volume of the tear reservoir. These results emphasise the necessity to include systematic examination of Tear Meniscus Height as part of a complete ocular surface assessment in glaucoma patients for early diagnosis and treatment of tear film abnormalities.

One of the aims of this study was to determine if there was a significant difference in level of tear production in glaucoma subjects living with and without diabetic mellitus in Owerri. An obvious statistically significant difference in mean tear production comparing the two groups (p<0.001) was observed. Moreover, the Chi-square analysis revealed a statistically significant connection between diabetes status and the grade of tear production (χ2 = 42.086, *p <* .*001*), suggesting a considerable impact of diabetes mellitus on tear production among the subjects with glaucoma. These data indicate that diabetes exerts an extra detrimental effect on lacrimal gland function beyond the ocular surface abnormalities previously seen in glaucoma and its long term medical therapy.

A statistically significant difference (p < 0.001) observed in the present investigation is similar to the findings, which observed strong relationships link between decreased tear secretion and the progression of diabetic retinopathy, longer duration of diabetes and poor glycaemic control (p<0.05).^31^ This similar findings in both studies suggests that diabetes mellitus plays an important role in lacrimal gland function and contributes to tear deficiency.

Extended duration of topical glaucoma medication, polypharmacy and formulations containing preservatives have been identified as important indicators of dry eye disease (p<0.05) in glaucoma patients.^32^ A multi-variable study showed that these factors influenced the probability of ocular surface disease separately.

Despite diabetes not being one of the focus variables in the study, the findings support the current study in that patients are particularly vulnerable to tear film abnormalities, which can be aggravated by the presence of diabetes. Similar results were reported by a study that observed considerable deterioration in ocular surface characteristics in glaucoma patients using topical medicines. The authors found that OSDI scores were worse (p = 0.008), and there was significant changes in Schirmer test scores (p=0.009).^29^ These findings confirm the present investigation in showing that glaucoma treatment impairs the ocular surface health, and diabetes exacerbates the reduction of tear production.

Results of the present study are also supported by the systematic review and meta-analysis which reported considerably reduced Schirmer I test in diabetes group values compared with non-diabetic controls. This single meta-analysis estimated total impact size spanning 59 studies, indicating a decrease in tear production in persons with diabetes mellitus.^33^

The outcomes of this study lend general support to strong evidence that diabetes mellitus considerably exacerbates tear production deficiency in glaucoma patients. The most important results taken from the independent samples t-test and the Chi-square analysis reveal that diabetes affects not only the mean rate of tear production, but also its clinical classification. These findings emphasize the need for regular tear function evaluation in glaucoma patients living with diabetes inorder to initiate early therapies to prevent ocular surface problems, enhance adherence to therapy and to maintain vision.

Using Independent sample t-test, statistically significant difference was observed for mean tear meniscus height in the two groups (*p=.034*). The Chi-square analysis further demonstrated statistically significant correlation of diabetes status with categorical grading of tear meniscus height (χ2 = 6.680, p = .010). These results suggest that diabetes status was related to the continuous measure and clinical grading of the tear meniscus height among the glaucoma subjects living with diabetes mellitus. The mean tear meniscus height in glaucoma subjects living with diabetes mellitus was 0.8 ± 0.3 mm, compared with 0.7 ± 0.3 mm in glaucoma subjects living without diabetes. While the difference between the two means was small, the *p-value of* .*034* suggests that the difference was still significant at the 0.05 level of significance. Likewise, the notable chi-square result shows that the distribution of glaucoma subjects according to tear meniscus height varied in term of diabetes status.

A previous study which compared glaucoma controls and patients with no glaucoma reported a substantial difference was seen in TMH of the medically managed glaucoma patients and age-matched controls. Patients with medically treated glaucoma had significantly lower mean TMH than controls (*p < 0.001*). Tear meniscus height was likewise considerably decreased in glaucoma group (p<0.001) whereas patients on combination treatment had larger decreases in tear meniscus height than single drug treatment.^34^ Although the difference in that direction was not the same as the present study, both studies showed that the tear meniscus height differ widely among glaucoma groups. In the present investigation, two groups who both had glaucoma, but varied in their diabetic status were compared. Thus, the chronic glaucoma treatment may have decreased tear volume in both groups in the present investigation, whereas diabetes may also have impacted drainage of tears, reflex secretion or clearing of the tears.

The recent findings might as well be explained by a study that investigated alterations in the ocular surface with asymptomatic glaucoma patients who have been on long-term topical antiglaucoma medicines, patients who had trabeculectomy, and healthy controls. The study showed that medical management of glaucoma causes a significant degradation of various ocular surface parameters in glaucoma patients compared to the other groups (*p < 0.05*).^39^ Tear meniscus height was not evaluated as a categorical outcome and no independent samples t-test or Chi-square test was done but their statistically significant results corroborate the present study which shows a significant difference in tear meniscus height between glaucoma subjects living and without diabetes mellitus, utilizing independent samples t-test and Chi-square test. Thus, both studies show that persistent glaucoma treatment contributes to alterations in the ocular surface, moreso, with possible additional impact on tear dynamics by diabetes mellitus in the present study.

The research works used in comparison to this study did not demonstrate Chi-square statistics for TMH grading. In these studies compared, TMH was evaluated as a continuous variable using tests of difference of means or longitudinal difference. Therefore, the provided *p-values* were compared to the independent samples t-test result of the current study but not directly with its Chi-square statistic. The Chi-square result of the current study, χ^2^ = 6.680, *p = 0.010*, adds more evidence that diabetes status was also linked to the category grading of TMH. Thus diabetes affected not just the mean TMH value but also the distribution of subjects over the given TMH grading.

This observed discrepancy may be attributed to the impact of diabetes on the functioning of lacrimal unit. Chronic hyperglycemia may be responsible for corneal neuropathy, autonomic dysfunction, ocular surface inflammation and microvascular damage. These alterations can interfere with reflex tearing, basal secretion, blinking and tear drainage. In people with glaucoma who have been exposed to chronic topical medicines and preservatives, diabetes may hence, lead to complicated changes in tear volume. This may emerge as reduced tear volume in a group of persons or as increased tear meniscus in some others, due to reflex tearing or poor tear drainage.

However, the statistically significant independent samples t-test result and Chi-square findings in this study indicated that diabetes status was related to changes in TMH among glaucoma subjects in Owerri. However, recent research using both groups as seen in this study are limited, and diabetes-specific studies demonstrate that diabetes substantially changes TMH, while glaucoma specific researches show that topical medication and therapy have considerable impact on the tear meniscus height. The moderately elevated TMH in glaucoma subjects living with diabetes in the present study may be thus considered as changes in tear dynamics and not necessarily due to better tear function. Tear meniscus height, should be considered in conjunction with Schirmer tear production, tear-film break-up time, ocular surface staining and discomforting symptoms in glaucoma subjects living with diabetes.

Major related published studies only compared diabetes subjects with non-diabetic controls or glaucoma patients on medical therapy to healthy or treatment-naïve controls. Hence, information from diabetes-specific and glaucoma-specific research was used to interpret the current conclusion, although their disparities in population and design are well acknowledged. Thus, a major finding is that this study addresses a gap in literature, which is, direct comparison of TMH in glaucoma subjects living with or without diabetes mellitus. Most of the published research considered just one of these conditions versus healthy controls.

The outcomes of this study lend general support to strong evidence that diabetes mellitus considerably exacerbates tear production deficiency in glaucoma patients. The most important results taken from the independent samples t-test and the Chi-square analysis reveal that diabetes affects not only the mean rate of tear production, but also its clinical classification. These findings emphasize the need for regular tear function evaluation in glaucoma patients living with diabetes inorder to initiate early therapies to prevent ocular surface problems, enhance adherence to therapy and to maintain vision.

## 5. Conclusion

Diabetes mellitus dramatically deteriorates the ocular surface function in glaucoma subjects by decreasing tear production, altering the tear meniscus height and increasing the severity of ocular surface symptoms. The substantial differences found in tear production and tear meniscus height suggest that diabetes mellitus worsens ocular surface dysfunction in glaucoma patients beyond the effects associated with glaucoma and long-term topical antiglaucoma therapy alone. Routine glaucoma care, especially in patients with diabetes mellitus, should include a full ocular surface evaluation including Schirmer I test, TBUT, TMH, and OSDI assessment to allow early detection and management of ocular surface disease, better treatment adherence, and improved visual outcomes.

## Data Availability Statement

The raw data supporting the findings of this study cannot be shared publicly due to institutional ethical restrictions regarding participant privacy.

## Acknowledgement

The authors acknowledge Prof. Emmanuel C. Esenwah for his immeasurable support and guidance through this study.

